# Characterising Negative Symptoms in Schizophrenia: CHANSS study protocol

**DOI:** 10.1101/2025.05.30.25328412

**Authors:** Noham Wolpe, Clàudia Aymerich, Ying Jin, Marta Martin-Subero, Paloma Fuentes-Perez, Claudia Ovejas-Catalan, Sara Salas-Rad, Renata Zirilli, Sophie Shatford, Rebecca Cox, Megan Cartier, Ana Catalan, Anna Mane, John Pratt, Lisa Airey, Paul Stanley, Adrianne Close, Andrew Hall, Javier Vazquez-Bourgon, Francesco Del Santo, Maria-Paz Garcia-Portilla, Nuria Segarra, Yi-Jie Zhao, Paul C Fletcher, Masud Husain, Peter B Jones, Emilio Fernandez-Egea

**Author notes:** Corresponding author: Emilio Fernandez-Egea MD PhD, Department of Psychiatry University of Cambridge, 128 Tenison road CB1 2DP Cambridge, UK.

## Abstract

Negative symptoms in schizophrenia, particularly motivational deficits, pose significant challenges to treatment and recovery. Despite their profound impact on functional outcomes, these symptoms remain poorly understood and inadequately addressed by current interventions. The CHANSS (Characterising Negative Symptoms in Schizophrenia) study aims to dissect the cognitive mechanisms underlying motivational impairments by focusing on three interconnected domains: executive cognition, motivational cognition, and meta-cognition. This large, international, cross-sectional study recruits a heterogeneous sample of patients across illness stages (from first-episode psychosis to treatment-resistant schizophrenia) and uses a comprehensive cognitive battery, clinical scales, self-report measures, and computerised cognitive tasks. Four novel tasks assess key processes in motivated behaviour: option generation, reward-based decision-making, risk sensitivity, and performance self-evaluation. By incorporating control for secondary influences like depression, psychosis, sedation, and illness chronicity, the study seeks to identify distinct cognitive and behavioural subtypes within motivational dysfunction. CHANSS tests the hypothesis that specific patient profiles exhibit predominant impairments in one or more cognitive domains, which may differentially affect goal-directed behaviour. The study design allows exploration of hierarchical relationships between cognitive processes, such as how executive deficits may cascade to impair motivation and self-evaluation. Ultimately, CHANSS aims to advance mechanistic understanding of motivational deficits in schizophrenia and pave the way for personalised, targeted interventions. Its findings may inform future clinical trials and contribute to a shift away from one-size-fits-all approaches toward more effective, stratified treatment strategies in schizophrenia.

## Introduction

Schizophrenia is a chronic and disabling psychiatric disorder affecting approximately 1% of the global population and remains a leading cause of disability worldwide [1]. Although considerable research efforts have focused on the pathophysiology and treatment of psychosis, negative symptoms have historically received less attention despite their significant impact on patients’ long-term outcomes [2]. They include anhedonia, avolition, asociality, blunted affect, and alogia, frequently prove resistant to both pharmacological and psychosocial interventions, substantially impairing patients’ social and occupational functioning [3].

Negative symptoms have been conceptualized to have two principal dimensions: diminished emotional expression (blunted affect and alogia) and deficits in motivation and pleasure (avolition, anhedonia, and asociality) [4–6]. Emerging evidence suggests these dimensions are psychometrically and potentially biologically distinct [7, 8]. Controversially, much of the research into negative symptoms has approached them as a unitary construct, possibly impeding the development of effective therapeutic strategies. A multifaceted approach, where each dimension is studied independently, may enhance our understanding of their distinct mechanisms. Given its critical role in determining functional outcomes, we focus specifically on motivation.

Motivational deficits in schizophrenia can be conceptualised as a syndrome involving multiple components, but distinguishing causal factors from confounders has been challenging. Despite diverse underlying mechanisms, these deficits converge on a common observable endpoint: impaired or absent goal-directed behaviour. For consistency with terminology used in other clinical fields and in neuroscience, we use the term ‘apathy’ to refer to the observable reduction in goal-directed behaviour [9]. Deconstructing apathy in schizophrenia into its specific cognitive components can help to delineate precise deficits, potentially deepening our understanding of schizophrenia’s complex symptomatology and guiding targeted interventions [10, 11].

The possible mechanisms underlying apathy in schizophrenia include executive dysfunction, motivational processing deficits and maladaptive beliefs [11]. Conceptually, these explanations impact different cognitive levels, which we categorise as executive cognition, motivational cognition and meta-cognition. First, executive cognition refers to the capacity to plan, initiate, and sustain goal-directed actions [12], which is strongly associated with apathy in schizophrenia [13]. Executive cognitive relies on foundational cognitive processes such as attention and working memory, and these functions are commonly assessed using standard neuro-cognitive assessments batteries, such as the Brief Assessment of Cognition [14] and MATRICS [15]. Second, motivational cognition refers to the specific cognitive processes related to the decision of whether to engage in goal-directed actions, and if so, which action to choose. The decision requires the valuation of effort and reward [16], and has been shown to be altered in schizophrenia [16, 17]. Motivational cognition has been assessed using decision-making paradigms [18, 19]. Third, meta-cognition encompasses the ability to monitor and evaluate one’s own performance; in schizophrenia, this is often characterised by defeatist beliefs and low self-efficacy, which can further inhibit motivation [20].

These three cognitive processes are likely interrelated and partially dependent on one another, which would make it difficult to separate their independent contribution to apathy in schizophrenia. However, the relative contributions of executive, motivational, and meta-cognitive processes to apathy in individuals with schizophrenia. Clarifying these contributions is essential for developing targeted interventions and refining existing therapeutic approaches. Moreover, clarifying which clinical factors are associated with which cognitive process is further crucial for adapting treatment. For example, secondary causes of negative symptoms, such as sedation related to antipsychotic medication [8], depression, disorganisation, psychosis and social stigma [21], can all contribute differentially to these cognitive processes.

Similarly, illness stage has been shown to differentially influence motivational cognitive: Two studies employing the same effort-based decision-making task in patients with schizophrenia found distinct results in patient populations at different illness stages. Specifically, individuals with chronic schizophrenia are less inclined to accept offers involving high rewards regardless of effort [19]. By contrast, individuals in earlier stages of the illness were less inclined to accept offers demanding high effort regardless of reward [18], indicating potentially different meta-cognitive contributions. Together, it is important to measure the three cognitive processes alongside key clinical variables relevant for apathy.

The CHANSS (Characterising Negative Symptoms in Schizophrenia) project aims to investigate the cognitive processes underlying apathy in schizophrenia, while measuring common secondary factors affecting apathy. To this end, the project will utilise a large, multi-site, multicultural and heterogeneous sample of individuals diagnosed with schizophrenia at various illness stages, from first-episode psychosis to treatment-resistant schizophrenia (TRS). This diversity enhances the generalisability of findings and allows exploration of how the cognitive processes relevant to apathy vary across the illness spectrum [22]. The principal aim of the project is to identify different subtypes of motivational impairments in patients based on the relative contribution of the three cognitive processes underlying apathy. The study employs an extensive battery of psychometric and cognitive assessments, alongside computer-based tasks designed to measure executive, motivational, and meta-cognitive aspects of goal-directed behaviour (Fig. 1).

**Figure 1.**
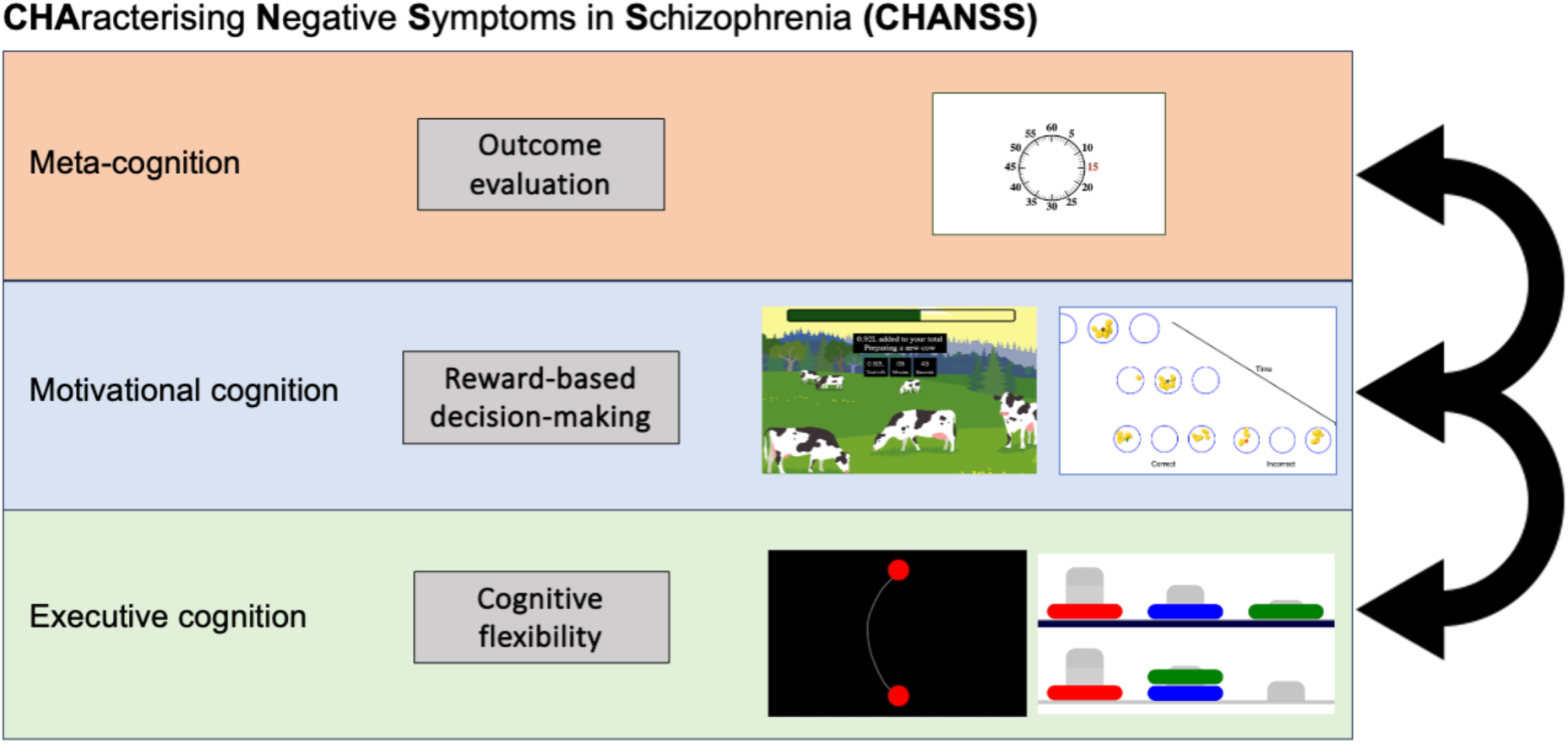
Schematic of theoretical and experimental framework. The project Characterising Negative Symptoms in Schizophrenia (CHANSS) aims to assess each of the three key cognitive processes hypothesised to underlie apathy in schizophrenia. It will quantify their relative contribution to apathy and their differential associations with clinical variables relevant to apathy, such as illness stage and depression.

General hypotheses:

- Group differences will emerge across all tasks, with patients performing less optimally than controls on tasks related to the three cognitive components.
- Some of these differences might be due to potentially treatable secondary causes of poor motivation (e.g., depression or drug-induced sedation). These secondary factors will selectively impair specific components within the motivational syndrome.
- Patients can be grouped into clusters or subtypes according to behavioural differences and relative contribution of the three (executive, motivation and meta-) cognitive components.
- A hierarchical effect will be observed, whereby dysfunction in more basic cognitive functions (e.g., executive dysfunction) would disrupt higher level processes (e.g., motivational and meta-cognition).

## Materials and Methods

### Study Design

The CHANSS (Characterising Negative Symptoms in Schizophrenia) study is a multicentre, experimental, cross-sectional study designed to investigate the cognitive underpinnings of motivation in schizophrenia. The study employs a comprehensive battery of clinical assessments, cognitive tasks and self-reported measures to evaluate motivational deficits across different illness stages, including first-episode psychosis and treatment-resistant schizophrenia (TRS).

Recruitment dates: Recruitment started in May 2021 and is expected to continue until July 2025.

### Study Sites

The CHANSS study was conducted across multiple international research centres, including academic institutions and mental health centres in the United Kingdom (Cambridge, London, Humber, Doncaster), Spain (Barcelona, Bilbao, Oviedo and Santander) and China (Shanghai). In the UK, the study benefits from the support of the National Institute for Health and Care Research Clinical Research Network (NIHR-CRN), which facilitates participant recruitment, site coordination, and data management.

### Inclusion and Exclusion Criteria

Inclusion Criteria:

– Age between 18 and 65 years.
– Diagnosis of schizophrenia according to the International Classification of Diseases, 10th Revision (ICD-10) criteria, with more than one year of diagnosis.
– Stable antipsychotic medication regimen for at least six weeks prior to study enrolment.
– Capacity to provide informed consent.

Exclusion Criteria:

– Presence of neurological disorders or significant medical conditions that may affect cognitive functioning.
– History of traumatic brain injury with loss of consciousness exceeding five minutes.
– Current substance use disorder (excluding nicotine) within the past six months.
– Intellectual disability (IQ < 70).
– Use of anticholinergic medication, except for hyoscine in cases of clozapine-induced sialorrhea.

### Ethical Considerations

The study protocol has been reviewed and approved by the relevant Institutional Review Boards (IRBs) and Ethics Committees at each participating site. In the UK, the protocol received ethical approval from the Health Research Authority (HRA; IRAS:295622; 21/WA/0056) and was registered with the NIHR Clinical Research Network Portfolio. This protocol included the other sites, but each site obtained their local ethical approval. The study complies with the principles outlined in the Declaration of Helsinki and Good Clinical Practice (GCP) guidelines. Written informed consent is obtained from all participants prior to any study-related procedures. Data confidentiality and participant privacy are maintained throughout the study in accordance with the General Data Protection Regulation (GDPR).

### Coordination and Validation

To ensure consistency and data quality across sites, standardised training sessions and inter-rater reliability assessments were conducted to validate clinical scales and ensure consistency in data collection. This process includes video-based training and cross-site workshops facilitated by experts in psychopathology assessment. The same clinical research forms (CRF) were used in all sites and each site had language-specific scales and tasks.

### Study Procedure

Participants undergo a comprehensive assessment protocol, including clinical interviews, cognitive tasks, and self-report questionnaires. The assessment is conducted over one or two sessions, depending on participant preference and tolerance, each lasting approximately 2-3 hours.

1. Screening and Informed Consent: Participants are screened for eligibility based on inclusion and exclusion criteria. Informed consent is obtained prior to any assessments.
2. Clinical Assessments: Administration of standardised clinical rating scales by trained researchers.
3. Cognitive Tasks: Participants complete a battery of computerised tasks designed to assess various components of motivation.
4. Self-Report Measures: Participants complete questionnaires to evaluate subjective experiences related to motivation and negative symptoms.

### Clinical Assessments

– Positive and Negative Syndrome Scale (PANSS): A clinician-rated scale with 30 items assessing positive symptoms, negative symptoms, and general psychopathology. It uses a 7-point Likert scale to evaluate symptom severity [23].
– Brief Negative Symptom Scale (BNSS): A 13-item clinician-rated scale designed to assess five negative symptom domains: blunted affect, alogia, anhedonia, asociality, and avolition. Each item is rated on a 7-point scale [24].
– Calgary Depression Scale for Schizophrenia (CDSS): A 9-item clinician-administered scale focusing on depressive symptoms specific to schizophrenia, differentiating them from negative or extrapyramidal symptoms [25].
– Personal and Social Performance Scale (PSP): A clinician-rated tool assessing social functioning across four domains: socially useful activities, personal and social relationships, self-care, and disturbing and aggressive behaviours [26].
– Brief Assessment of Cognition in Schizophrenia (BACS): A 20-minute validated cognitive battery to measure functions such as working memory, attention, and verbal fluency [14].

### Self-Report Measures

– Apathy Motivation Index (AMI): A 18-item self-report questionnaire evaluating motivation across different life areas [27].
– Autism Spectrum Quotient (AQ-50): A self-report measure to screen for autistic traits that may overlap with negative symptoms [28].
– Apathy Evaluation Scale (AES): A self-rated scale with 18 items designed to measure apathy, covering behavioural, cognitive and emotional domains [29].

### Cognitive and Behavioural Tasks

The study incorporates four computerised tasks designed to investigate the three cognitive processes underlying motivated behaviour (see above). These tasks, although novel to this population, have been validated in healthy individuals. The tasks were designed to be less taxing in terms of possible fatigue and to be 5 to 15 minute long to facilitate engagement. Participants are seated comfortably, interacting with a computer via keyboard, mouse or touchscreen, with clear instructions and breaks provided to minimise fatigue. All tasks were programmed in JavaScript and run on a dedicated server with Just Another Tool for Online Studies for backend [30].

#### 1. Option Generation Task

Objective: To assess cognitive flexibility, creativity and action fluency we will deploy a simple, established task that has shown a strong correlation with self-reported apathy in healthy people [31], Lack of ability to generate options for behaviour may be particularly relevant to negative symptoms in schizophrenia [32]. Along with the BACS, this task will test executive cognitive aspects of the motivational syndrome in schizophrenia.

Task description: Participants use a stylus pen to draw paths between two red circles on a touchscreen tablet (Fig. 2). All sites used tablets of the same make and model (Samsung Galaxy Tab S6 Lite). One red circle appeared at the top and one at the bottom of the display. In the first control condition, participants were asked to draw as many paths as possible between the bottom and top red circle for 30 seconds. In the main experimental condition, participants were asked to draw as many *different* paths as possible between the red circles. Drawn paths appeared on the screen as they were drawn and remained visible throughout the task to reduce working memory demands. Participants were free to create curved or straight paths, and the paths could overlap or intersect. To ensure paths were counted as valid, they had to start at the bottom circle and end at the top circle.

**Figure. 2.**
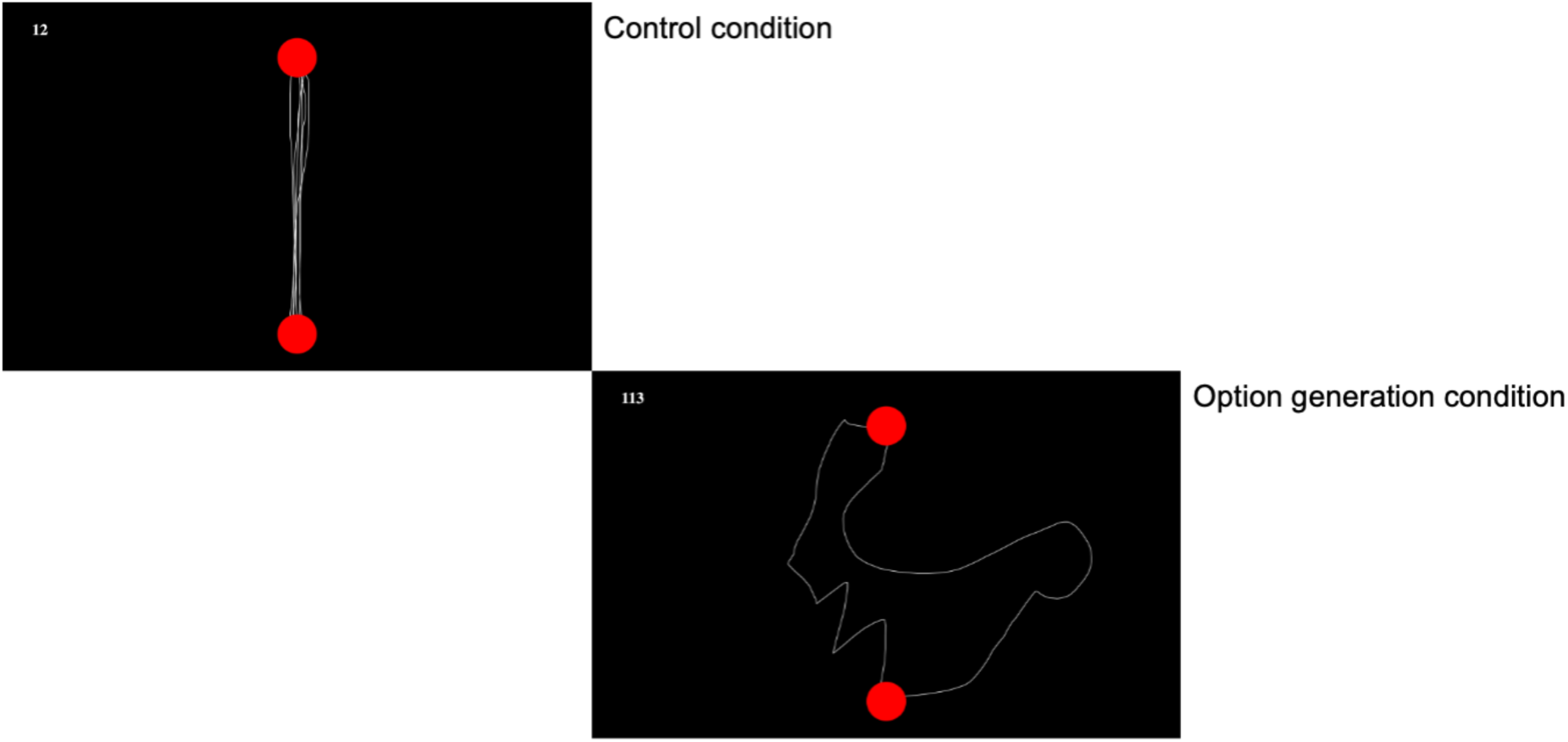
Illustration of the option generation task.

Cognitive process: The control condition accounts for individual differences in movement speed. The main experimental condition measures fluency (the total number of paths generated) and uniqueness (the distinctiveness of each path relative to others). Uniqueness will be computed by quantifying the geometric (Euclidean) distance between paths, as done previously [31], while accounting for basic differences in movement speed. This approach provides an objective measurement of creative output and flexibility, essential for understanding goal-directed behaviour.

General hypothesis: This task has not been previously applied in schizophrenia research. However, the task has been used in people with Parkinson’s disease [31] and with depression [33]. The hypothesis is that patients will show reduced fluency and reduced uniqueness, and, and these would be related to executive dysfunction in patients, as measured using the BACS.

#### 2. Milkman Task

Objective: To operationalise the decision to switch in a time-constrained paradigm, as part of the motivational cognitive process. Specifically, the task examines reward-based decision-making in a foraging framework, evaluating tendencies to persist with diminishing returns—a behaviour hypothesised to be altered in people with affective conditions and apathy [34, 35] and to be dependent on dopamine [36]

Task description: Participants are instructed to collect as much milk as possible over a 10-minute period (Fig. 3). To milk a cow, they hold down the spacebar, during which a bucket on the screen visually represents the accumulating milk. Participants are told that milking would become progressively harder over time. Milk accumulation follows an exponential decay function:

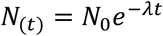

**Figure 3.**
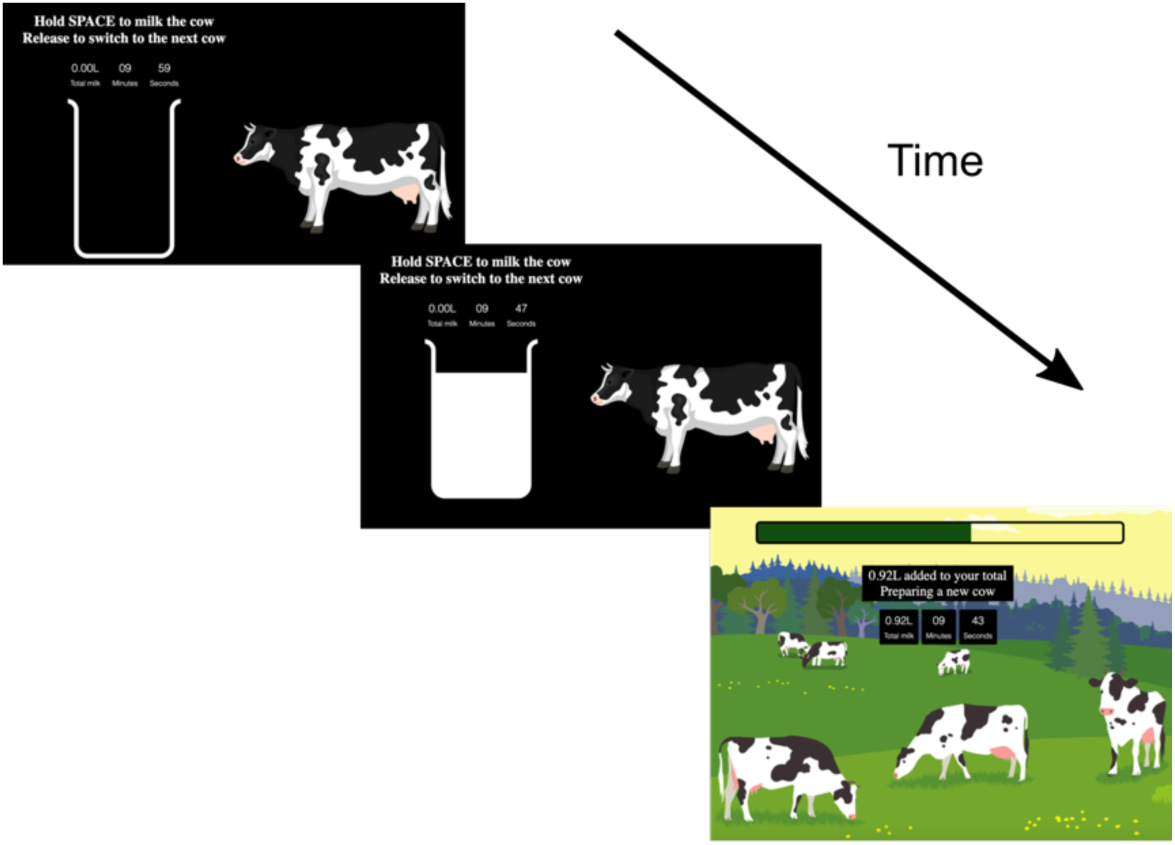
Schematic of foraging in the milkman task.

Where *N*_(t)_ is the instantaneous accumulation rate at time *t*, *N*_0_ is the initial rate, and λ is the rate of decline. There are four cow types, created by crossing two initial reward rates (0.01 or 0.02) with two decay rates (2e-4 or 4e-4), resulting in a 2×2 design: High-Slow, High-Fast, Low-Slow, and Low-Fast. These conditions are designed to vary in overall profitability and optimal harvesting duration and are presented in a pseudorandom sequence such that every four-trial cycle includes one of each cow type.

When participants release the spacebar, they enter a fixed 4-second travel period representing the time needed to reach the next cow. During this phase, a progress bar shows the remaining travel time, and the total milk earned so far is updated. Pressing the spacebar prematurely during the travel screen triggers an on-screen warning lasting two seconds. A countdown timer displays the remaining time for the task, and participants’ cumulative milk total is always visible. The task begins with a 1-minute practice session to familiarise participants with the interface and response requirements.

Cognitive process: The task assesses decision-making related to switching behaviours. The cognitive process involves cost-benefit analysis and temporal discounting, designed and analysed using a foraging framework. The primary outcomes will be ‘stay duration’ and the related ‘exit threshold’ which is the reward rate at the decision to switch on each trial. Based on these, we will compute an ‘offset’ parameter, reflecting participant’s overall tendency to stay longer or shorter than the optimal policy; and a ‘scaling’ parameter, reflecting how much participants exaggerate differences between conditions compared to the optimal policy [37].

General hypothesis: This task has not been previously applied in schizophrenia research. We have recently validated it in healthy participants in an online study [37]. Other foraging tasks have been used in the context of affective disorders [35], Attention Deficit Hyperactivity Disorder [38], and Parkinson’s disease [36]. It is hypothesised that motivational symptoms will be associated with overstaying behaviour, reflecting a failure to adapt to diminishing returns.

#### 3. Tokens Task

Objective: To examine decision-making under uncertainty and failure aversion, using a variant of a task from evidence accumulation literature [39], as part of the motivational cognitive process.

Task description: Participants completed a decision-making task in which they observe the movement of tokens from a central circle to two lateral circles and are asked to make a decision whether the left or right circle will have the most tokens by the end of the trial (Fig. 4). At the start of each trial, 15 tokens and a central cross are presented in a central circle. These tokens move incrementally, one at a time, towards either the left or right circle at pre-defined intervals. The movement pattern is determined by a pre-generated binary sequence, ensuring variability across trials while maintaining an equal number of left– and right-winning trials. The winning side was set to receive 8 or 9 tokens, ensuring that trials are not overly predictable.

**Figure 4.**
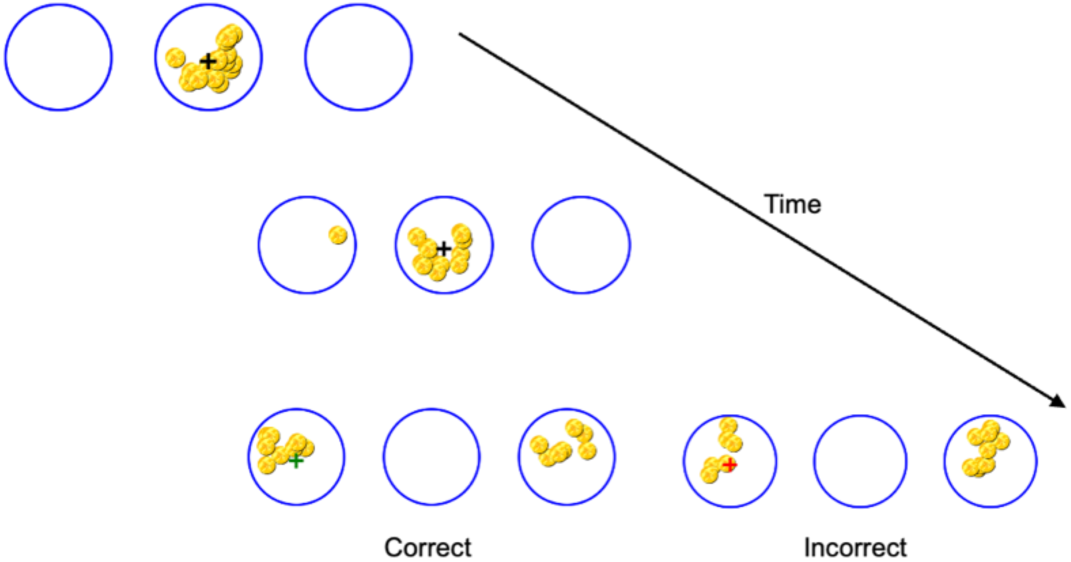
Schematic of decision-making process in the tokens task.

Participants are instructed to make a decision as early as they felt, using the left or right arrow key to indicate their response. Once a decision is made, the cross moves to the chosen circle, and the remaining tokens are quickly distributed from the middle circle. If the choice is correct, the cursor turned green, a positive auditory cue is played, and the text ‘X number of tokens earned’ appears in green, where X is the number of tokens earned on that trial. If incorrect, the cursor turns red, a negative auditory cue is played, and the text ‘no tokens earned’ appears in red. If participants do not make a choice before all tokens had moved, a ‘timeout’ warning appears, and participants receive incorrect feedback (red circles and negative auditory cue).

Participants earn the number of tokens remaining in the middle circle when a correct decision is made. This speed accuracy trade-off is emphasised in the instructions given to participants, while they are encouraged to collect as many points as possible. Unlike previous studies [40], we chose not to add explicit penalty for incorrect choices and focussed on the effect of implicit loss feedback. Each participant completes 100 trials, with an equal number of left– and right-winning trials presented in a randomised order.

Cognitive process: The task evaluates the decision when and which action to choose. It measures evidence accumulation, decision thresholds and risk-taking. Main outcome measures were number of tokens drawn until decision and response (‘right’ or ‘left’). We will fit a computational model to compute each participant’s loss aversion, with the hypothesis that people with schizophrenia will require more evidence before committing to a choice due loss aversion, over and above general slowness.

General hypothesis: This task has not been previously applied in schizophrenia research. It is expected to reveal altered risk sensitivity, providing a novel perspective on cognitive biases in this population.

#### 4. Clock Stopping Task (Performance Beliefs Task)

Objective: To explore meta-cognitive deficits in schizophrenia, particularly defeatist performance beliefs, which suggest that patients generalise negative thoughts about their abilities, which leads them to avoid goal-directed behaviours [20, 41].

### Task Description

The Performance Beliefs Task (Fig. 5) involves participants estimating their ability to stop a moving clock hand at a target location (‘time’). The task has two phases: First, during the action (hand stopping) phase, participants observe a moving red clock hand that rotated clockwise or counterclockwise and are asked to stop it as close as possible to a predesignated target time (marked in colour red). They press the spacebar to stop the red clock hand. Second, after stopping the clock hand, during the estimation phase, the clock hand disappears, and the target time turns black again. Participants are prompted to estimate where they stopped the red hand by manually adjusting a second blue clock hand using the arrow keys. The experiment consists of 50 trials, with pseudorandomised hand movement directions and stopping targets. Participants are also given a time limit for making their estimates. If they respond too late, they receive a ‘timeout’ message and repeat the trial.

**Figure 5.**
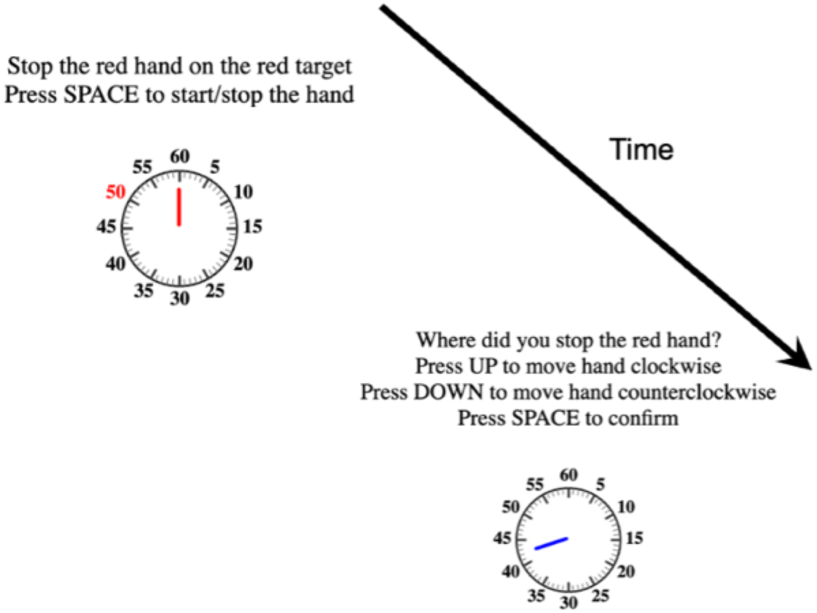
Schematic of meta-cognitive beliefs in the clock stopping task.

Cognitive process: The task was designed to assess meta-cognitive beliefs about performance, testing how well participants could predict and evaluate their own actions. Each trial has two main outcome measures: Performance error, calculated as the difference between the target position and where the red hand was stopped, and estimation error, calculated as the difference between their estimated and actual stopping position. These measures can be fit using a Bayesian model that assumes that belief about the stopping position combines noisy sensory input with prior expectations, reflecting cognitive biases in performance estimation [42].

General hypothesis: This task has not been previously applied in schizophrenia research. Similar paradigms have been used in Parkinson’s disease and healthy populations [43, 44], We hypothesise that patients will exhibit diminished expectation of success, compared to the typical optimistic expectations people show in the task [42].

### Power Calculation

The study was designed to have >90% power to detect correlations with medium effect size (r=0.3) at the level of ɑ = 0.05 independently for each task. To this end, we aimed to recruit at least 112 patients. To account for multiple comparisons (which will be corrected using False Discovery Rate method), and considering the expected high group heterogeneity, we aimed for a large (n > 130) sample size.

### Data Management

All data are collected using secure, encrypted electronic data capture systems. Data is pseudo-anonymised on each site, assigning a unique identifier (e.g., CC001) to each volunteer. Data are stored in compliance with local regulations and specifically local data protection regulations. Quality control measures include regular audits, and inter-rater reliability assessments for clinical ratings.

## Discussion

Along with cognitive deficits, negative symptoms are the most pervasive in schizophrenia, determining long-term prognosis and jeopardising functional recovery [45–47]. However, the advances in treating psychotic symptoms have not been mirrored in improving motivation in schizophrenia. There are different culprits for this lack of advance, including the limitation of the current scales and the amalgamation of apathy with emotional expression into the unitary concept of negative symptoms. We have previously highlighted the difficulties in the current scales as well as the need for focusing on either motivation or emotional deficits when studying their biology and when testing new treatments [48, 49].

This large, multicentre, multicultural study aims to identify the cognitive and clinical factors underlying the motivational syndrome in schizophrenia. It integrates three leading hypotheses of apathy: that it arises from executive dysfunction, impaired motivational decision-making, or meta-cognitive deficits such as defeatist beliefs. The study investigates whether distinct profiles of these deficits exist and how they interact. While we acknowledge that executive, motivational, and meta-cognitive processes are interrelated, we argue that quantifying their relative contributions in individual patients is essential—particularly given the high heterogeneity of schizophrenia. Such profiling could inform personalised treatment strategies, moving beyond the broadly applied pharmacological interventions that have shown limited success to date [50]. For example, individuals with prominent defeatist beliefs may respond better to cognitive behavioural therapy, whereas those with executive impairments may benefit more from cognitive remediation. Furthermore, by accounting for secondary factors that influence motivational deficits—such as depression or medication side effects—we may identify more immediate, practical interventions, such as antidepressant use or antipsychotic dose adjustments. These insights can help tailor clinical management to the needs of individual patients and lay the groundwork for stratified treatment approaches.

In sum, CHANSS study offers a chance to disentangle the complex syndrome of poor motivation, in order to enable implementable interventions and informing future mechanism-led clinical trials.

## Disclosures

Dr. Aymerich is supported by the Alicia Koplowitz Foundation and has received personal fees or grants from Janssen Cilag and Neuraxpharm, outside the current work. Dr. Catalan reports having received support to attend scientific meetings from Janssen, ROVI, and Lundbeck in the last five years. She is also supported by the Instituto de Salud Carlos III, Spanish Ministry of Economy and Competitiveness. Dr. Mane has received support to attend meetings or served as a advisor or speaker for Janssen Cilag, Neuraxpharm, Otsuka, Lundbeck and Angellini. Dr. Vazquez-Bourgon has received financial support to attend scientific meetings or as advisor or speaker at educational events from Janssen, Lundbeck and Angellini Pharma.

M.H. is a consultant for NeuHealth. EFE has received consultancy honoraria from Boehringer-Ingelheim (2022), Atheneum (2022) and Rovi (2022-24), speaker fees by Adamed (2022-24), Otsuka (2023) and Viatris (2024) and training and research material from Merz (2020) and editorial honoraria from Spanish Society of Psychiatry and Mental Health (2023-). EFE is also deputy editor of the British Journal of Psychiatry, but has not role in the evaluation of this paper. Rest of authors reports no conflicts of interest with this project.

## Funding

NW was supported by an Israel Science Foundation Personal Research Grant (1603/22) and NSF-BSF-NIH Computational Neuroscience (CRCNS) grant (2024628). Dr. Vazquez-Bourgon is supported by ISCIII (PI20/01279) and Plan Nacional sobre Drogas (2021/079; EXP2022/08898). NS is supported by NIHR Greenshoots program. PCF is funded by provided by the Bernard Wolfe Health Neuroscience Fund and a Wellcome Trust Investigator Award to PCF (Reference No. 206368/Z/17/Z). M.H. re supported by the Wellcome Trust and the NIHR Oxford Health Biomedical Research Centre. PBJ is supported by NIHR (PGfAR 0616-20003) and Wellcome, and is co-founder of Cambridge Adaptive Testing ltd. Dr Fernandez-Egea is supported by the 2022 MRC/NIHR CARP award (MR/W029987/1), specifically to this project. All research at the Department of Psychiatry in the University of Cambridge is supported by the NIHR Cambridge Biomedical Research Centre (NIHR203312) and the NIHR Applied Research Collaboration East of England. The views expressed are those of the author(s) and not necessarily those of the NIHR or the Department of Health and Social Care.

## Data Availability

All data produced in the present study are available upon reasonable request to the authors

